# Evaluation of OCT biomarker changes in treatment-naive neovascular AMD using a deep semantic segmentation algorithm

**DOI:** 10.1101/2022.06.16.22276342

**Authors:** Ben Asani, Olle Holmberg, Johannes B Schiefelbein, Michael Hafner, Tina Herold, Hannah Spitzer, Jakob Siedlecki, Christoph Kern, Karsten U. Kortuem, Amit Frishberg, Fabian J. Theis, Siegfried G Priglinger

**Author notes:** **Corresponding Authors:** Fabian Theis & Siegfried G. Priglinger, MD, FEBO, Department of Ophthalmology, Ludwig-Maximilians-University Munich, Mathildenstrasse 8, 80336 Munich, Germany. Authors contributed equally to this work (Ben Asani, Olle Holmberg, Johannes B Schiefelbein, Fabian J. Theis, Siegfried G Priglinger).

## Abstract

**Purpose:** To determine real life quantitative changes in OCT biomarkers in a large set of treatment naive patients undergoing anti-VEGF therapy. For this purpose, we devised a novel deep learning based semantic segmentation algorithm providing, to the best of our knowledge, the first benchmark results for automatic segmentation of 11 OCT features including biomarkers that are in line with the latest consensus nomenclature of the AAO for age-related macular degeneration (AMD).

**Design:** Retrospective study.

**Participants:** Segmentation algorithm training set of 458 volume scans as well as single scans from 363 treatment naive patients for the analysis.

**Methods:** Training of a Deep U-net based semantic segmentation ensemble algorithm leveraging multiple deep convolutional neural networks for state of the art semantic segmentation performance as well as analyzing OCT features prior to, after 3 and 12 months of anti-VEGF therapy.

**Main outcome measures:** F1 score for the segmentation efficiency and the quantified volumes of 11 OCT features.

**Results:** The segmentation algorithm achieved high F1 scores of almost 1.0 for neurosensory retina and subretinal fluid on a separate hold out test set with unseen patients. The algorithm performed worse for subretinal hyperreflective material and fibrovascular PED, on par with drusenoid PED and better in segmenting fibrosis. In the evaluation of treatment naive OCT scans, significant changes occurred for intraretinal fluid (mean: 0.03µm^3^ to 0.01µm^3^, p<0.001), subretinal fluid (0.08µm^3^ to 0.01µm^3^, p<0.001), subretinal hyperreflective material (0.02µm^3^ to 0.01µm^3^, p<0.001), fibrovascular PED (0.12µm^3^ to 0.09µm^3^, p=0.02) and central retinal thickness C0 (225.78µm^3^ to 169.40µm^3^).The amounts of intraretinal fluid, fibrovascular PED and ERM were predictive of poor outcome.

**Conclusions:** The segmentation algorithm allows efficient volumetric analysis of OCT scans. Anti-VEGF therapy provokes most potent changes in the first 3 months and afterwards only acts as a stabilizing agent. Furthermore, a gradual loss of RPE hints at a progressing decline of visual acuity even beyond month 12. Additional research is required to understand how these accurate OCT predictions can be leveraged for a personalized therapy regimen.

**Précis:** Novel high performance segmentation algorithm shows most volumetric changes under anti-VEGF therapy in oct biomarkers occur in the first 3 months. Afterwards the injections seem only to serve as a stabilizing agent.

## Introduction

Vascular retinal diseases represent a heterogeneous field with multiple etiologies. In many countries age related macular degeneration (AMD) and diabetic retinopathy are the leading causes of blindness and are therefore of high socioeconomic importance^1,2,3^. Western lifestyle and the aging population are expected to contribute to a strong increase in the incidence of these diseases^4^.

While there are only limited therapeutic options for the dry form of AMD (dAMD), the pharmacological inhibition of the vascular endothelial growth factor (VEGF) has revolutionized treatment of neovascular AMD (nAMD) in the past two decades and has therefore become the gold standard in treatment. The intravitreal injection of anti-VEGF agents such as Ranibizumab (Lucentis®), Aflibercept (Eylea®), Bevacizumab (Avastin®) and the just newly approved Brolucizumab (Bevou®) targets the VEGF-guided growth of pathological choroidal nevovascularisations that would normally lead to excessive intraretinal and subretinal fluid or excessive subretinal hemorrhages and lastly to the development of fibrotic tissue or geographic atrophy leading to high visual defects and central scotomas. While large pivotal studies for Lucentis® (MARINA-Study) and Eylea® (VIEW-Study) showed a visual acuity gain of 7.2 to 9.3 EDTRS letters after one to two years^5,6^, follow up studies observed a mean loss in visual acuity of 8.6 ETDRS letters after 7 years^7^. The major cause for this seems to be the development of retinal fibrosis in the context of a wound healing reaction that is shown to occur in approximately 50% of all patients after two years^8^. Apparently, a big part of the scar consists of fibrotically changed vascular membranes due to anti-VEGF therapy^9,10^. In addition to the limitations mentioned above, the high therapy intensity of the established anti-VEGF agents with monthly to bimonthly injections over several years poses a major challenge for both patients and the health care system. A recent study by Chopa et al shows an 11-fold increase in annual intravitreal injection from 2009 to 2019 and is projected to continue to rise^11^.

Besides fibrosis there are several other OCT biomarkers that seem to play an important role in determining the course of disease, the progression into advanced stages and response to treatment^12–15^. Generally, the influence or the importance of OCT biomarkers for visual acuity progression or progression into fibrosis in neovascular AMD is not fully understood. For example, recent studies have shown that ignoring subretinal fluid (<200µm at the foveal center) does not change outcome in visual acuity for patients under anti-VEGF therapy but can lessen their treatment burden^16^. Other studies by Christenbury et al and Dhrami-Gavazi et al have shown that a relatively stable fibrovascular pigment epithelial detachment may correlate with protection for the development of macular atrophy^17,18^. This shows that new findings in further analyzing morphologic features in these patients can give new insights to “older” treatment regimens. To understand changes in these features a basic descriptive understanding of biomarkers in treatment naive patients in a real life setting is crucial. Study conditions can lead to “selection bias” (i.e. through inclusion criteria) and can not offer the same insight into the variety of patients or their compliance as do real field studies. Manually analyzing these treatment naive biomarkers and their changes over time becomes difficult and time consuming once we want to study a greater set of patients. Additionally, the variety of morphologic features in AMD add much to the complexity of the scan and generate high volume data making the manual process close to impossible. Hence, the use of automatic segmentation can offer a great deal of help in analyzing, describing and understanding the changes of the OCT biomarkers as well as their influence on the overall therapeutic outcome. In this respect, deep learning algorithms have already been shown to successfully automate tasks such as disease classification of color fundus photographs and retinal OCT images, a key non-invasive imaging modality in Ophthalmology, as well as support medical decision making in many other medical fields ^19–23^. Further, in the field of deep semantic segmentation, the task of classifying individual pixels in an image with numerous applications in medicine, many advances have been made in both natural scenes as well as biomedical applications ^24–27^. In this project we propose a deep learning based semantic segmentation algorithm trained with 458 manually annotated macular OCT scans, to allow automatic segmentation, and thus automatic labeling, and volumetric analysis of clinical features of a large series of treatment naive patients eyes undergoing anti-VEGF treatment. It furthermore allows us to give a one-of-a-kind extensive description on the distribution of morphologic features as well as disease biomarkers of that patient group in a real life setting. To the best of our knowledge, we are the first to present an extensive AMD segmentation algorithm including disease biomarkers and other common morphological OCT features as well as validate this on independent test sets of previously unseen patients combined with detailed analysis of the inter-annotator variance for ambiguous and hard to annotate features.

## Methods

This case series included patients with newly diagnosed treatment naive neovascular age related macular degeneration in the study eye and had a follow up observational period of at least three and twelve months. Treatment naive was defined as never having had any form of intravitreal injection (anti-VEGF or other) in our institution or elsewhere. The first intravitreal injection was received at time of diagnosis. Exclusion criteria were comorbidities such as central retinal vein occlusion, retinal branch occlusion, diabetic macular edema, uveitis and other conditions that can lead to the development of intra-or subretinal fluid such as macular telangiectasia, central serous chorioretinopathy and intravitreal injection of any other drug than anti-VEGF. The study was approved by the institutional review board of our institution and adhered to the tenets of the Declaration of Helsinki. Written informed consent was obtained from each participant prior to the intervention and all testing outlined herein.

### Treatment regimen

Patients received an upload of three monthly injections of any of the anti-VEGF agents (Ranibizumab, Aflibercept or Bevacizumab) and were then treated according to the Treat and Extend regimen: They were either extended for two weeks or continued on a monthly injection routine^28^.

### Patient identification

To identify patients suffering from a neovascular AMD, we queried our data warehouse for all patients receiving intravitreal Injections of anti-VEGF between 2013/03/11 and 2020/07/09. Diagnosis of neovascular AMD was confirmed after proof of choroidal neovascularization in initial Fluorescein angiography. We interpolated the data set to 3 measurement points: Start date is the time of the first intravitreal injection. Next monitoring point is after 3 months and lastly after 12 months of treatment.

### Preoperative examinations

Examinations before intravitreal injections included testing best corrected visual acuity using standard ETDRS chart at testing distance of 4 meters, intraocular pressure using non contact tonometry, dilated indirect fundoscopy as well as spectral-domain optical coherence tomography of the macula (Spectralis; Heidelberg Engineering GmbH, Heidelberg, Germany). The metrics “counting fingers”, “hand movement” were converted to 1.98 and 2.28 logMar respectively as previously described by Lange C et al and Schulze-Bonsel et al^29,30^. There were no patients included with the metric “light perception”. All visual acuity values in this study are given in logMAR units.

### Segmentation data sets

To create the segmentation algorithm, a set of 458 macular OCT scans, each from a different patient, were annotated by ophthalmological residents with a fellowship in medical retina (B.A., J.B.S. and M.H.) using the annotation tool LabelMe^31^. They were then validated by three retinal experts (J.S., C.K. and T.H.) and re-labeled in case of any discrepancies. Each fellow was assigned his or her own set, roughly a third of the 458 scans, randomly. In this process, we followed the Consensus Nomenclature for Reporting Neovascular Age-Related Macular Degeneration of the AAO (American Academy of Ophthalmology) for disease biomarkers and used 11 different OCT labels^32^ also including other common morphological features giving us the possibility to analyze the scan as a whole and not exclusively for the biomarkers. The annotation was made pixel wise, i.e. each pixel in the image was assigned one of the 11 classes: Epiretinal Membrane (ERM), Neurosensory Retina (NR), Retinal pigment epithelium (RPE), Intraretinal fluid (IRF), Subretinal fluid (SRF), Subretinal hyperreflective material (SHRM), Drusenoid pigment epithelial detachment (DPED or Drusenoid PED), Fibrovascular pigment epithelial detachment (FPED or Fibrovascular PED), Fibrosis, Choroid, Posterior hyaloid membrane (PHM). For annotation examples as well as exact class distribution statistics see supplement. Further the class camera effect and the vitreous cavity were automatically labeled. For camera effect this was derived as completely black regions in the OCT images and for vitreous cavity as background above the neurosensory retina. The central retinal thickness C0 was defined as the quantification (in µm^3^) of the labeled neurosensory retina in that region according to the ETDRS grid^33^.

An additional data set of 30 scans were sampled and then annotated to measure the inter annotator variation between the three annotators for a selection of ambiguous features. To quantify feature ambiguity as well as establishing an upper bound for how well these features can be expected to be segmented, the ophthalmological fellows all annotated the same scans. After doing so, a consensus annotation was produced by a panel including the retinal experts by either annotating a new scan or selecting one of the annotated scans from the annotators. In total, each of the 30 scans in the second data set had three annotations from the three fellows in medical retina as well as a fourth annotation produced by consensus voting.

### Model architecture

The model used is a deep ensemble^34^ of networks, for increased generalization in the presence of limited number of samples^35^, for segmentation. Each network in the ensemble is the same, just trained with different random weight initializations. The architecture used for the models in the ensemble is described below.

The algorithm used for segmenting the retinal OCT scans is a deep convolutional neural network (CNN)^36^ of a U-net type architecture^24^. Specifically the network consists of eleven convolution layers followed by batch normalization^37^ and relu activation functions^38^. The convolutional layers use padding so as to not alter the dimensions of the feature maps and have kernel size set to three throughout the network. Each convolutional layer is initialized using the He normal initialization^39^ at the start of the training. In the encoder, every two convolutional layers are followed by a max pooling operation making a total of five max poolings, reducing the resolution size of the input from 256 to 8 for the feature maps. Here, the original images are linearly resized from 512 to 256 pixels heigh and width. The first convolutional layer is set to have 64 filters and this number is doubled after every max pooling layer yielding a maximum of 1024 filters in the bottleneck of the architecture. The number of filters are then halved after every transposed convolution in the decoder. Between the encoder and the decoder a dropout layer with probability 0.2 is applied for regularization. In the decoder, transposed convolutions as well as two layered convolutional blocks as described above are applied consecutively until the original input dimension is reached. Finally, a convolutional layer with kernel size one and a softmax activation function is applied to achieve the final output of the network.

### Model training

The models were trained using the Adam optimizer using the categorical cross entropy loss, with an initial learning rate of 0.001, found to be optimal through hyper parameter tuning on a validation set. In total, each network was trained for 2000 epochs. During training, random flipping, rotation, brightness and gaussian noise, with a scale of (2.5, 12.75), were used to augment the data. Each with a probability of 0.2 of being applied to an image or image and mask, except for rotation which was assigned a probability of 0.5 of occurring. The images and annotation masks were split into a train, validation and test set consisting of 338, 84 and 36 images respectively. Further, an ensemble of networks was created for each image in the 36 image test set using a leave-one-out validation scheme and adding the remaining test images to the training data set. In total five models were trained for each test image resulting in 180 different models. At inference time, the softmax outputs for each class and pixel, from all five models, were averaged to obtain an ensemble prediction. The class with the highest average softmax score yielded the final prediction. For the inter doctor variance data set the 10 models, out of 180, with the best validation scores were selected to form an ensemble from which the predictions were obtained in the same way as for the test set.

### Model evaluation

The segmentation model is evaluated using the F1 score, i.e. the harmonic mean between precision and recall, a standard evaluation metric for semantic segmentation tasks. The score is then presented for each class. Further the inter doctor variance is presented as the F1 score between each annotator retinal expert, also called annotator and the 10 model ensemble against the consensus annotation. As all pixels are concatenated for all images, as typically done in semantic segmentation tasks, no standard deviation metrics between images are provided. The statistically evaluated model was then used for automatic segmentation of 18522 OCT scans from 378 eyes enabling the statistical analysis of morphological OCT features including nAMD biomarker distribution on treatment naive patients and how they are affected by anti-VEGF injections.

### Statistical analysis

All statistical analysis was performed using the python programming language with the scipy stats software package^40^. Normality of data was assumed due to sufficiently large sample sizes well above 30 as specified by the central limit theorem^41^. We applied the independent samples t-test for parametric comparisons. The level of statistical significance was defined as p<0.05.

The code for the models and training procedures as well as result analysis will be made available through the public Github repository upon publication.

## Results

### Deep learning segmentations accurately quantifies presence of clinical features in retinal OCT images

The segmentation algorithm was trained using 338 doctor-annotated OCT scans from AMD patients to infer the volumes of 11 different clinical retinal features (See **Methods**). The segmentation algorithm segmented the clinical features with a top performance of 0.98 F1-score for features neurosensory retina and subretinal fluid. The lowest F1-scores were observed for features epiretinal membrane, drusenoid PED and subretinal hyperreflective material (see Fig. 1a). Test set examples of the segmented features can be seen in figure 1b, showing the variety of features accurately segmented in unseen patients. The annotators, on the other end, highly agreed on subretinal hyperreflective material, fibrovascular PED and drusenoid PED, while largely disagreeing in the case of fibrosis (see Fig. 1c). Overall, the segmentation algorithm performed worse than the annotators on subretinal hyperreflective material and fibrovascular PED, on par with respect to drusenoid PED and better than all annotators when segmenting fibrosis (see Fig. 1c). In figure 1d we see examples of consensus, annotator and segmentation algorithm predicted segmentations for example OCT images. One can clearly see how fibrosis is annotated as fibrovascular PED by annotators II and III and while detected, subretinal hyperreflective material is overall segmented differently by the algorithm than the annotators and consensus segmentations.

**Figure 1:**
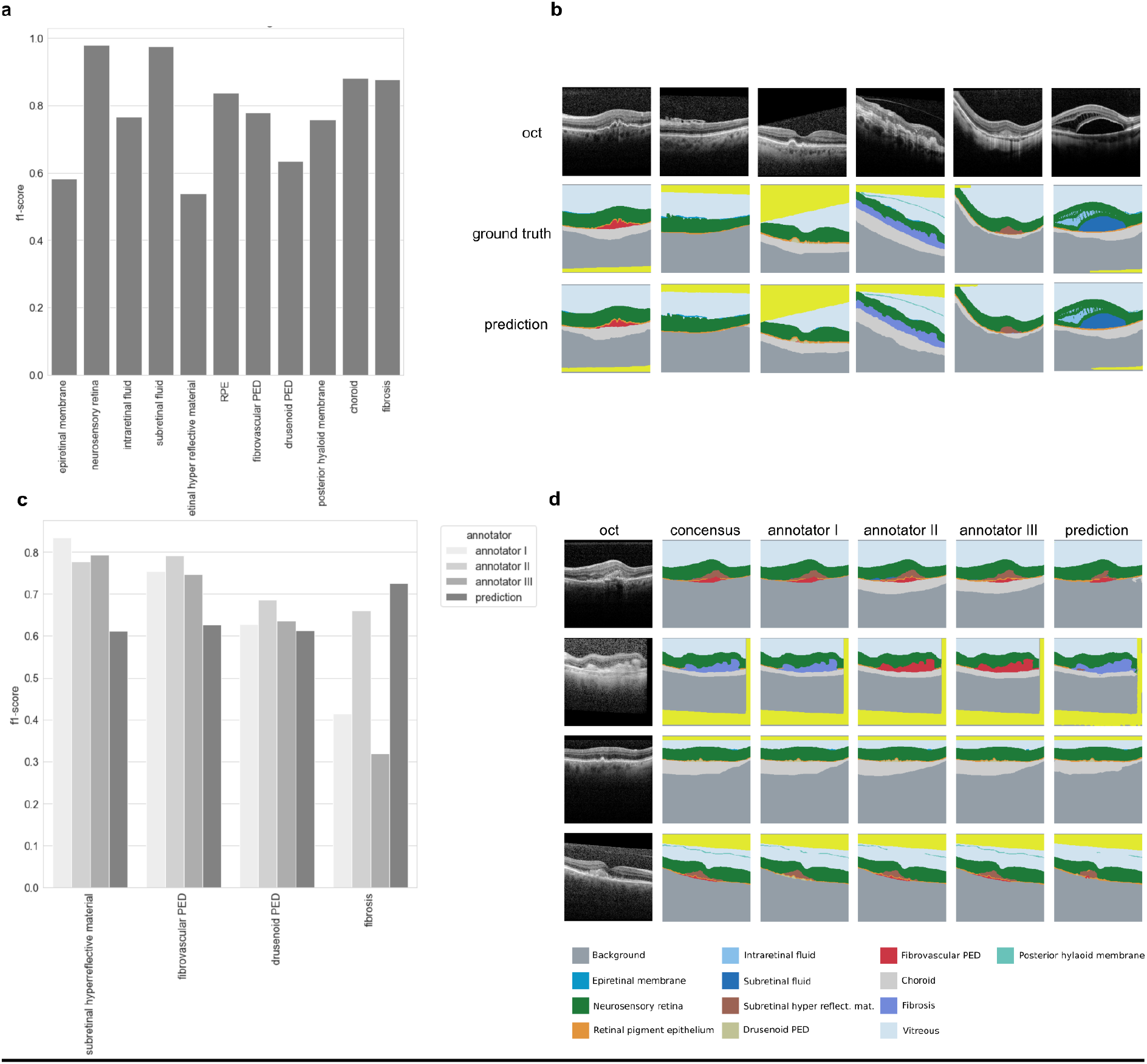
Deep learning segments clinical features on par with human experts from retinal OCT images. (a) F1 scores for 11 clinical features segmented on a test set from 37 to the algorithm previously unseen patients. (b) Example of OCT images selected to illustrate various segmentation classes with ground truth and predicted segmentation maps (c) F1 scores for subretinal hyperreflective material, fibrovascular PED, drusenoid PED as well as fibrosis on 30 challenging test patients containing these features. (d) Example OCT images with consensus ground truth, annotations from three different annotators as well as predicted segmentation maps displaying segmentation of multiple features.

### Most morphological changes of treatment naive patients occur during the first three months of anti-VEGF therapy

Through our data warehouse we identified 592 eyes of 536 patients, out of 1656 AMD patients in total, that were treatment naive and had the necessary follow up visits to fit the study. After filtering for the inclusion criteria as described in the methods sections (i.e. follow-up time) a total of 378 eyes consisting of 18522 OCT scans from 339 different patients were segmented and analyzed. Of those patients 144 were male and 234 female with an average age of 82 ± 8 years. Pathological biomarkers in treatment naive patients that were most prominent on initial presentation were by far fibrovascular pigment epithelial detachment (Mean: 0.12 µm^3^, SD: 0.19 µm^3^), followed by subretinal fluid (Mean: 0.08 µm^3^, SD: 0.26 µm^3^), intraretinal fluid (Mean 0.03 µm^3^, SD 0.08 µm^3^) and subretinal hyperreflective material (Mean: 0.02 µm^3^, SD: 0.05 µm^3^, see Fig 2). The mean central retinal thickness C0 (CRT) was 225.78 µm with SD 90.36 µm in the cohort of treatment naive patients. After 3 and 12 months of intravitreal treatment the CRT decreased compared to baseline (3 months: mean 169.40 µm, SD 64.50 µm, p-value < 0.001; 12 months: mean 169.42 µm, SD 59.03 µm, p-value < 0.001) which is mostly related to the resolution of fluids. Mean number of injections was 3.8 ± 1.5 after 3 months and 8.3 ± 3.5 after 12 months, meaning that on average the therapy regimen was extended at some point during the observed time frame. After 3 months of intravitreal treatment there was little but significant change in fibrovascular PED from 0.12 µm down to 0.09 µm (p-value = 0.02) but no difference afterwards (mean 12 months: 0.10, p-value from month 3-9: 0.62). The overall change of this marker over the course of twelve months was barely not statistically significant (p-value: 0.05) meaning that therapy in general did not lead to real regression of the fibrovascular PED over a long term treatment regimen but rather contributes to a stabilization of this biomarker.

**Figure 2.**
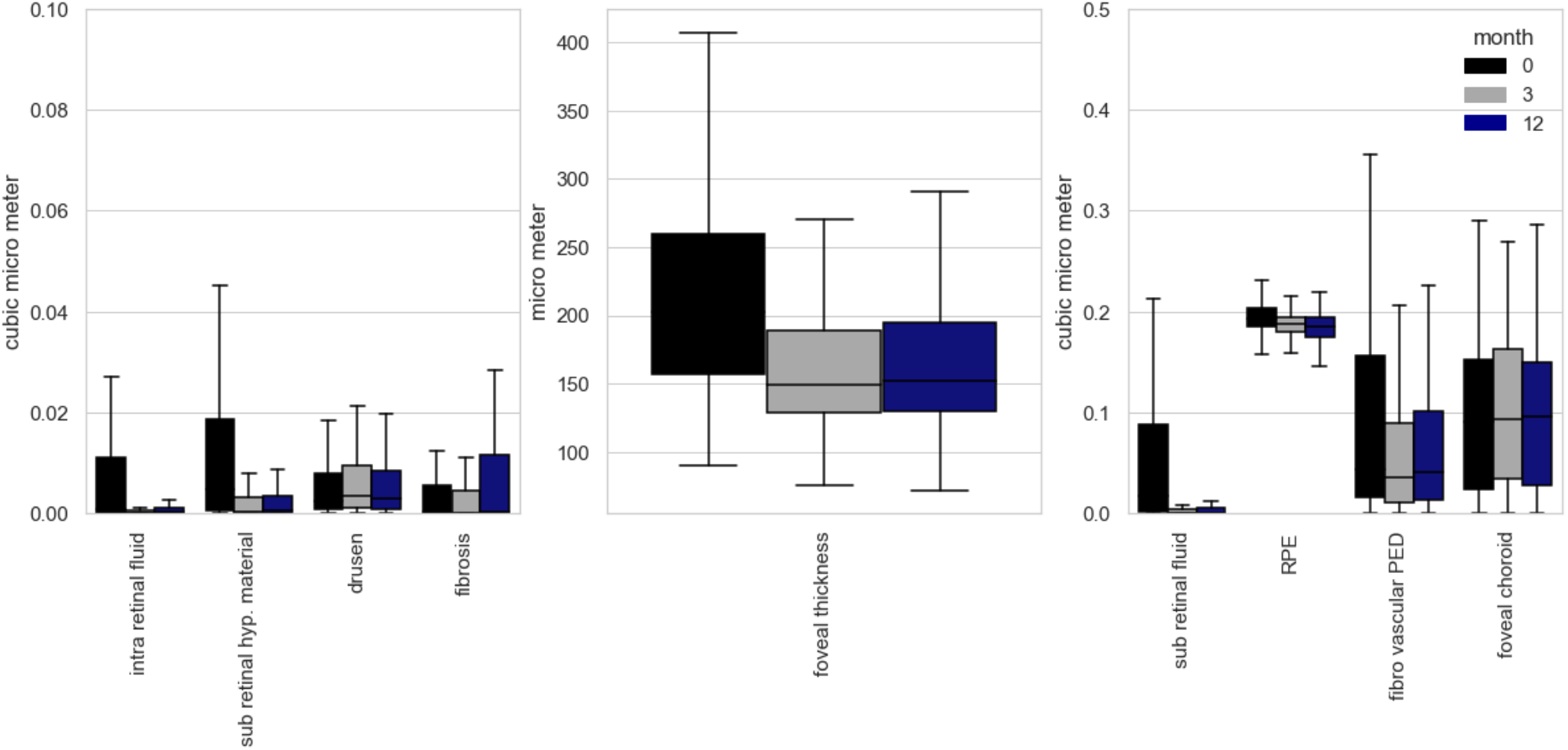
Volumetric changes of treatment naive OCT biomarkers under anti-VEGF treatment. Significant reduction was seen in IRF, SRF, SHRM, RPE and central foveal thickness.

Difference in means as tested by t-tests showed a statistical significant change in IRF (p < 0.001), SRF (p < 0.001), SRHM (p < 0.001) and CRT C0 (p < 0.001) after 3 months of anti-VEGF therapy but no significant improvement or change after that initial upload of 3 injections until month 12 occured, meaning setbacks under therapy were uncommon and no further improvement could be reached. This was on par with the observed changes in visual acuity. Significant improvement was achieved from before treatment during the first 3 months (mean VA on month 1: 0.59 logMAR, mean VA on month 3: 0.50 logMAR; p-value: 0.003) but no significant change occurred afterwards up until month 12. The distribution of these and the other morphological features including disease biomarker amongst treatment naive patients is summarized in Figure 2 and Table 1. The RPE feature changed from initially 0.192 µm^3^ to 0.185 µm^3^ after 3 months (p < 0.01) and lastly 0.181 µm^3^ after 12 months (p-value month 3-12: 0.01) meaning there was a significant loss of RPE therefore hinting at a steady increase in atrophy. However, the cause-relationship between intravitreal injections and atrophy remains unclear as the resulting loss of RPE could be attributed to a natural disease progression. There was no change in choroidal thickness in 12 months even though there was a gradual loss in RPE.

**Table 1.** Mean volumetric values before (mean 1), three months (mean 3) and twelve months after initiation of anti-VEGF therapy. All values are in µm^3^.

|  | erm | irf | srf | srhm | rpe | fvped | drusen | phm | choroid | fibrosis | crt |
| --- | --- | --- | --- | --- | --- | --- | --- | --- | --- | --- | --- |
| mean 1 | 0.01 | 0.03 | 0.08 | 0.02 | 0.19 | 0.12 | 0.01 | 0.02 | 0.93 | 0.02 | 223.82 |
| mean 3 | 0.01 | 0.01 | 0.01 | 0.01 | 0.19 | 0.09 | 0.01 | 0.02 | 0.96 | 0.02 | 169.65 |
| mean 12 | 0.01 | 0.01 | 0.01 | 0.00 | 0.18 | 0.09 | 0.01 | 0.02 | 0.95 | 0.03 | 172.62 |
| sd 1 | 0.02 | 0.08 | 0.25 | 0.05 | 0.02 | 0.18 | 0.01 | 0.03 | 0.41 | 0.06 | 88.29 |
| sd 3 | 0.02 | 0.07 | 0.09 | 0.02 | 0.02 | 0.15 | 0.01 | 0.03 | 0.41 | 0.06 | 64.11 |
| sd 12 | 0.02 | 0.04 | 0.04 | 0.01 | 0.02 | 0.15 | 0.01 | 0.03 | 0.41 | 0.08 | 63.76 |
| p-value (1-3) | 0.62 | 0.00 | 0.00 | 0.00 | 0.00 | 0.02 | 0.44 | 0.39 | 0.33 | 0.59 | 0.00 |
| p-value (1-12) | 0.38 | 0.00 | 0.00 | 0.00 | 0.00 | 0.05 | 0.82 | 0.24 | 0.50 | 0.15 | 0.00 |
| p-value (3-12) | 0.70 | 0.94 | 0.86 | 0.32 | 0.01 | 0.59 | 0.30 | 0.74 | 0.76 | 0.05 | 0.52 |

We additionally used linear regression to model the interdependence between the different biomarkers and visual acuity outcome in 12 months. In treatment naive patients there was a negative correlation (meaning more of the feature was correlated with poor visual acuity outcome) between visual acuity and intraretinal fluid (p < 0.001), epiretinal membrane (p = 0.013), fibrovascular PED (p = 0.009) and fibrosis (p = 0.049). Since most of these features are most prominent in the active disease state (excluding epiretinal membrane) and bring immense structural changes to the retina, this finding does not deem very surprising. Interestingly, subretinal fluid had no correlation with a worse outcome in visual acuity, supporting the thesis of being protective against retinal atrophy and further decline of vision. Other findings are summarized in Table 2.

**Table 2.**
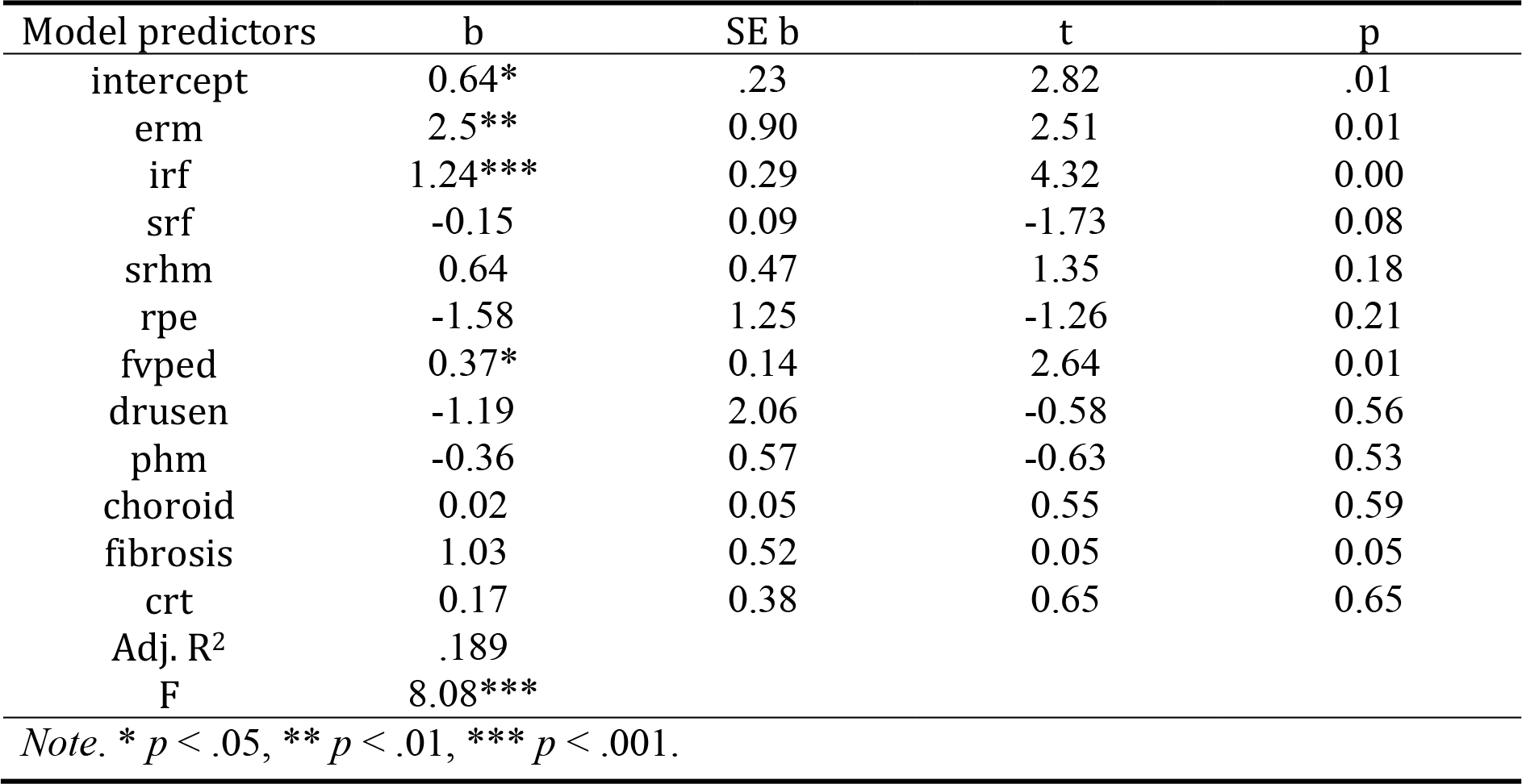
Results from multiple regression analysis of oct markers and their effect on 12 month visual acuity (n=336).

## Discussion

Structural changes and understanding of OCT images have been part of many major studies in the past. OCT imaging has changed AMD diagnostics and shaped the way patients are introduced into their treatment plans. To our knowledge, there is still missing an extensive morphologic description including disease biomarkers of never before treated nAMD patients. Most studies lack a full volumetric/quantitative interpretation of the images as manual segmentation seems unfeasible with high volume data. Therefore, in this study we introduced and showed the feasibility of a novel and high performing segmentation algorithm to present a thorough descriptive analysis of biomarkers in treatment naive nAMD patients in a real life setting and their overall plasticity after anti-VEGF treatment. Furthermore we identified several biomarkers that seem to contribute to a reduced prognosis in best corrected visual acuity after 12 months giving us a glance at predicting disease outcome.

### Contribution to the application of deep learning algorithms in understanding the Retina

Semantic segmentation of fluids, PEDs as well as retinal layers in retinal OCT images has been studied before^42–45^. To the best of our knowledge, this is the first project aiming to segment features as defined in the latest Consensus Nomenclature for Reporting Neovascular Age-Related Macular Degeneration of the AAO (American Academy of Ophthalmology) and uses 11 different OCT labels^32^. The only similar segmentation algorithm to our knowledge was developed by De Fauw and colleagues^22^ which used a similar segmentation network as a pre-processing method for predicting retinal disease. Our work differs with regards to two main aspects. First, we include the latest consensus nomenclature as specified by the AAO, and second, we provide an in-depth analysis of feature ambiguity by inter annotator variation analysis and test performance metrics reporting on a large number of unseen test patients. We show that segmentation of the above features is indeed possible to a high degree of accuracy (see Fig. 1a). While the algorithm segments at a similar proficiency as the annotators and even outperforms the retinal experts at segmenting fibrosis, we also clearly show the current limitation of segmentation of the retina by quantifying the ambiguity of the features fibrosis, drusen and subretinal hyperreflective material (see Fig. 2).

These features, which mainly occur in the subretinal region, can often, due to lower image quality, be hard to distinguish as they often share hyperreflective parts that can be mistaken for one another. These difficulties specifically present in transitions from fibrovascular PED to fibrosis as they would appear very ambiguous on OCT scans. For example, Ohayon et al segmented fibrovascular PED into three layers with a more hyperreflective layer 2 that did not respond as efficiently as the other layers following anti-VEGF treatment being suggestive of a fibrotic component in the PED^46^. Interestingly in accordance to that, the segmentation algorithm presented in this study occasionally segmented the fibrovascular PED into a fibrotic sub-compartment as well, making the segmentation process in this case potentially more accurate then the training set provided by the retinal fellows as they had to make a definite decision on which biomarker was present for the whole structure. However, this warrants further investigation. Lastly, due to the lower quality in some of the scans, it seemed difficult to precisely isolate the borders between the different OCT classes on a pixel-wise level in this OCT region. Quality issues in OCT-scans can have a variety of reasons and are a very common phenomena as already described by Domalpally et al^47^. For the purpose of this segmentation algorithm, it was crucial to be able to handle difficult images (quality-wise) as well, so we did not exclude scans with a lower image quality in the training sets.

Future work should aim to expand the number of features, such as adding Serous PED, segmented by the algorithm by collecting and annotating more data. Further, to deal with the difficulty of hard to annotate examples and annotator disagreement, future data sets should contain even more annotators and retinal experts annotating all the images in order to model the feature uncertainty.

### Distribution of Biomarkers and their overall plasticity on treatment naive patients

After proving the efficacy of the automatic deep segmentation algorithm, we described and examined the quantitative volumetric change over 12 months of therapy in OCT biomarkers in a large series of treatment naive nAMD patients after anti-VEGF therapy in a real life setting down to the microscopic level. We show that the most prominent change was to be seen in the first three months suggesting the initial upload phase of three injections as being the most potent for morphological changes to the retina. Interestingly, almost no changes could be determined afterwards until month 12, revealing that any further injection past the initial monthly upload phase only functioned as a stabilizing agent. The biomarkers most sensitive to change were usually the ones that have been most prominent at the beginning: IRF, SRF, SHRM, RPE and fibrovascular PED alongside CRT.

We showed that of those markers only intraretinal fluid and fibrovascular PED were predictive of a generally worse visual acuity outcome after 12 months. Although this seems logical, as higher amounts of those markers would indicate higher disease activity, we couldn’t find any prior evidence that would necessarily link the actual volumetric amount of fluid to a different outcome in contrast to fibrovascular PED for which there was prior evidence for example as described by Boyer et al^48^. One reason would be that patients with an abundance of these markers might have generally presented later after the first onset of symptoms and thus natural progression of the disease would have led to more irreversible retinal damages. Delay to treatment is a significant factor for poorer outcomes as previously described by Lim et al ^49^. Other than these, fibrosis significantly contributes to a worse outcome (which is again a marker of a progressed disease state) and interestingly epiretinal membrane. This is in direct contradiction to earlier findings by Alkin et al, where they concluded comparable results between patients with ERM and without ERM that were treated with bevacizumab^50^. However, reasons for our different findings could be a much higher number in investigated patients and the actual volumetric quantification of the observed epiretinal membranes in our oct scans. An increase of ERM did not necessarily mean more traction but it seems as simply a volumetric increase of this feature was enough to be predictive of a poorer outcome. It remains unclear why this would be the case. A possible explanation, at least in part, could be an increased traction over time (although in this work not possible to quantify) and thus more damage to the rpe and photoreceptor cells. However a missing increase in intraretinal fluid would contradict this. In additional contrast to Alkin et al. we did not solely include patients treated with bevacizumab, but rather conglomerated the different anti-VEGF injections alltogether. Although highly unlikely, this might be part of the reason for different findings in our work.

Similar descriptive studies have been done in the past but with a much smaller case series, manual segmentation and only some of the biomarkers segmented in this study which in general led to various findings^46,48,51,52^.

Lai et al observed treatment response for intraretinal cysts (IRC), subretinal fluid, pigment epithelial detachment and their correlation with BCVA changes for a time period of a year whilst setting similar time points as in our study (month 1, 3, 6 and 12)^51^. In 126 eyes only 33.3% showed a resolution of their PED whereas IRC resolved in 53.8% and SRF in 51.6% of cases^51^. In correlation with that we saw a statistically significant and prominent volumetric reduction in IRF and SRF, whilst the decrease in Fibrovascular PED was barely significant. This supports the findings of an actual lower resolution of PEDs that were observed in the aforementioned study. This could mean that while anti-VEGF may be sufficient in controlling the PED, it lacks strong efficacy in decreasing its size or making it disappear. Other observations by Golbaz et al demonstrated that intraretinal fluid and subretinal fluid immediately responded to anti-VEGF treatment albeit the overall plasticity of the morphological changes declined over time which surely corroborates with our findings. Interestingly the sub-RPE compartments showed the least or no changes to anti VEGF therapy^52^. The high immediate effect of anti-VEGF on especially IRF or SRF is all in all well documented in several other studies in the literature. Bolz et al for example showed a significant effect of anti-VEGF on retinal fluid compartments as early as one week after the injection whilst there was no significant change after the second and third injection ^53^. While this might be suggestive of cutting the loading dose to just one injection, the small number of cases (n=29) and missing long term observation of relapsing cases in that study warrants caution.

Our study is mainly limited by its retrospective nature. A true randomized, double blind prospective study would give us much more reliable information to what effect anti-VEGF actually influences all the OCT biomarkers but it would not be feasible to include controls groups as it warrants many ethical concerns. However, given the high number of real treatment naive patients in this study, the findings can more or less be very suggestive on the effect of anti-VEGF treatment. Moreover, it gives us a general idea on a broad population of patients, and what morphological properties they present upon first presentation. Other limitations obviously include the discussed feature ambiguities and the lower F1 scores for some of the biomarkers. While this could lead to some changes being misinterpreted, the F1 numbers are still quite high in general. In some cases they can be artificially lower as for certain features (especially membranes) it is difficult for manual annotators to segment correctly down to a pixel level with the precision that an automatic segmentation algorithm would. These micro-differences negatively impact F1 scores, albeit the algorithm being able to handle real segmentation perfectly.

Another limitation is that a steady decrease of RPE is not a real marker of “atrophy” or “geographic atrophy” and just gives us the idea or a hint towards such a feature. An adequate definition of atrophy in OCT volume scans is provided by Sadda et al where 3 criteria have to be met^54^: choroidal hyper transmission, attenuation of the RPE band, collapse or thinning of the outer retinal layer. A new training set for atrophy would be required that includes all three previously described properties to safely detect it as a biomarker for age-related macular degeneration.

To conclude, our study shows the feasibility of a high-performing segmentation algorithm, trained with a fixed set of 458 macular OCT scans, to quantitatively segment the whole region of a much larger number of OCT volume scans and thereby enable precise determination of volumetric changes down to a microscopic level. With that we were able to present a general description of the volumetric distribution of biomarkers in treatment naive patients and confirm that the most significant plasticity in biomarkers happens during the first 3 months of therapy with changes coming to a halt afterwards. For future work, this algorithm and findings could be used to analyze larger time series data sets and possibly predict the best possible therapy for each individual patient leading the way to a more personalized approach in treatment regimens.

## Data Availability

The code for the models and training procedures as well as result analysis will be made available through the public Github repository upon publication. All data produced in the present study are available upon reasonable request to the authors.

