## Supplement for "Evaluation of OCT biomarker changes in treatment-naive neovascular AMD using a deep semantic segmentation algorithm"

**Data, model training and automatic OCT segmentation:**

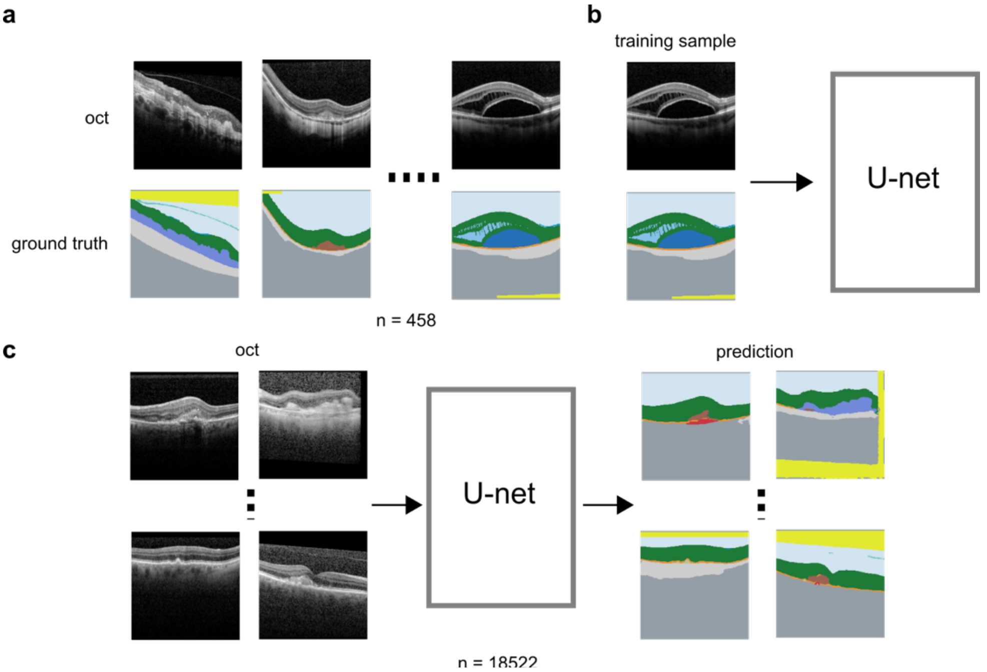


**Figure 3 Supplement.** *a, 458 OCT B-volumes are manually annotated for b, training U-net like deep segmentation models used for c, automatic segmentation of 18522 OCT volumes of longitudinal data.*

Our method relies on 458 manually annotated OCT B-scans for training, (b), of an ensemble of U-net like deep semantic segmentation models which are then used to automatically generate segmentation labels for clinical features of 18522 OCT B-scans from our data warehouse.

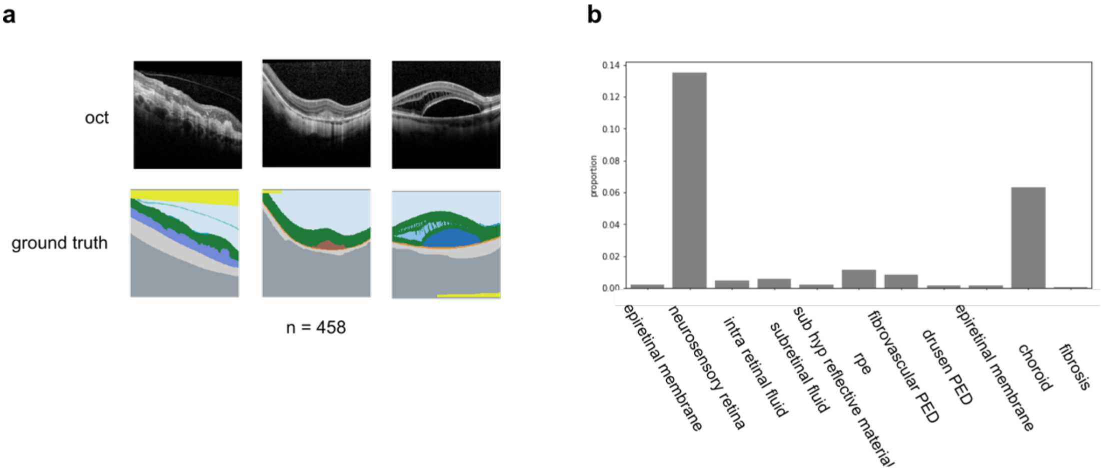


**Figure 4 Data Supplement.** a, three example OCT B-scans and their ground truth annotations. In b we see the proportion of each class label. Neurosensory retina and choroid dominates the labeled pixels.

In Figure 4 Data Supplement b the class imbalance between the clinical features annotated and later predicted in highlighted. Clear is here how neurosensory retina and choroid dominates the labeled pixels. Also rpe and fibrovascular PED are often annotation.
